# Designing to Implement Genomics Informed ASCVD Risk Assessment: Patient and Clinician Perspectives about Identifying and Managing the Underlying Causes of Severe Hypercholesterolemia

**DOI:** 10.64898/2026.08.10.26360146

**Authors:** Kelly M. Morgan, Gemme Campbell-Salome, Zachary M. Salvati, Muki Kunnmann, Dylan Cawley, Lauren Carr, Laura Ceballos, Samuel S. Gidding, Eimear E. Kenny, Amy R. Kontorovich, Tara Naib, Matthew T. Oetjens, Vikas Pejaver, Sabrina Suckiel, Matthew I. Tomey, Laney K. Jones, Miranda L.G. Hallquist

## Abstract

**Introduction:** Severe hypercholesterolemia has four primary causes: monogenic familial hypercholesterolemia (FH), polygenic hypercholesterolemia (PRS), severely elevated Lp(a) concentration, and hypercholesterolemia due to environmental/lifestyle/behavioral factors (i.e., no known genetic etiology). Here, we explore patient and clinician perspectives about the identification and management of each of these causes.

**Methods:** Patients with severe hypercholesterolemia with a primary language of English or Spanish and clinicians (primary care, genetic counseling, cardiology) across two health systems (Geisinger, Mount Sinai) participated in semi-structured interviews. Analysis was completed using an *a priori* codebook informed by Proctor’s implementation outcomes to identify themes influencing the identification and management of the underlying causes of severe hypercholesterolemia.

**Results:** A total of 28 patients and 25 clinicians participated. Patients emphasized the importance of receiving results directly from their clinician, requested take-home resources that mirrored the information from their clinician, were motivated to seek multidisciplinary care, and anticipated all results would be actionable, but that high-risk PRS and elevated Lp(a) may require more support (e.g., specialists, education) to act on. Clinicians stressed the importance of integrating workflows (e.g., test ordering) with the electronic health record, highlighted LDL-C levels and multidisciplinary care coordination as key to management, explained how they would tailor care to individual patients, and expressed a more limited understanding of Lp(a) and PRS result types based on their clinical experiences and, therefore, hesitation about the recommended clinical actions.

**Conclusions:** Patients and clinicians identified complementary determinants influencing the identification and management of the underlying cause of severe hypercholesterolemia. Participants welcomed risk information and requested a higher level of informational support and specialty expertise to appropriately manage high Lp(a) and PRS results. Integrating genomic information into risk assessments will require a partnership between general practitioners and specialists to provide a multidisciplinary approach to the identification and management of the underlying causes of severe hypercholesterolemia.

## Introduction

Severe hypercholesterolemia (LDL-C ≥ 190 mg/dl) requires lipid lowering therapy to prevent atherosclerotic cardiovascular disease (ASCVD), however it remains under-treated^1–4^. The causes of severe hypercholesterolemia include the presence of a familial hypercholesterolemia (FH) genetic variant, a high-risk polygenic risk score (PRS) for elevated LDL-C (i.e., high-risk PRS), extreme elevation of Lp(a), and environmental/lifestyle/behavior factors (i.e., no identified genetic cause)^5–7^. Previous work demonstrated that many cases were due to elevated Lp(a) (17.2%, 2,023/11,738), high-risk PRS (17.6%, 2,070/11,738) or a combination of elevated Lp(a) and high-risk PRS (7.8%, 913/11,738)^6^. Further, different etiologies predicted different future rates of ASCVD, suggesting that identifying the underlying cause of hypercholesterolemia may influence the intensity of LDL-C treatment^6^.

Testing for the underlying causes of severe hypercholesterolemia is currently at different stages of clinical implementation. FH testing has been clinically available and included in guidelines for over a decade, however most patients meeting clinical diagnostic criteria don’t receive genetic testing and testing is often limited to a specialty genetics setting^8,9^. More recently, published guidelines have recommended Lp(a) testing as a part of routine cardiovascular risk assessment, however Lp(a) testing has not been broadly adopted in routine care^10,11^. PRS testing is currently limited to the research setting, in the absence of clinical guidelines or widely available commercial testing options^12^. The evolving understanding of the genetic basis of severe hypercholesterolemia and emerging therapies pose challenges to clinicians’ understanding and preparedness to appropriately counsel patients^13^. It can be difficult for clinicians to differentiate the causes of severe hypercholesterolemia cases, particularly given the established role of environmental and lifestyle factors^14^. Further, limited patient understanding of this risk information and recommended management can impede adherence to treatment^15^. Incorporating newer identified causes (e.g., Lp(a) and PRS) may increase the overall complexity of counseling about risk assessment and management.

The aim of this study was to identify, among both patients and clinicians, barriers, facilitators, and recommended strategies to support the identification and management of the underlying causes of severe hypercholesterolemia (FH, high-risk PRS, elevated Lp(a), or environmental/lifestyle/behavioral factors). We applied a well-studied implementation science framework, the Conceptual Model of Implementation Research (CMIR)^16^, to examine the acceptability, appropriateness, adoption, and feasibility of integrating risk information based on the underlying cause of severe hypercholesterolemia into patient-centered care.

## Methods

This project is part of a larger multiple-method study, Improving Risk Stratification in Familial Hypercholesterolemia (RISK-FH, R01HL159182), which includes aims to build a population-based cohort to better stratify severe hypercholesterolemia based on underlying etiology and develop tailored ASCVD risk prediction. In-depth interviews were conducted across two health systems (Geisinger and Mount Sinai) to examine: 1) how clinicians would approach the diagnosis and management of severe hypercholesterolemia given the different underlying causes and 2) the needs and preferences of patients facing these results. Geisinger is a community-based learning health system serving approximately one million residents from central and northeastern Pennsylvania^17^. Across Geisinger’s coverage area, 35 counties are designated as rural. Mount Sinai is an integrated healthcare network based in New York City with eight hospital campuses and over 400 outpatient practice locations serving an ethnically and racially diverse patient population^18^. Both institutions have active genomic research and clinical cardiovascular genetic counseling, providing access to genetic testing for patients with severe hypercholesterolemia. This study was approved by the Geisinger Institutional Review Board (IRB #2021-0961), the Mount Sinai Institutional Review Board (IRB #23-00457) and follows the Standards for Reporting Qualitative Research (SRQR)^19^.

For patient interviews, adults with severe hypercholesterolemia were identified through clinician practices (A.R.K, M.I.T, T.N) at Mount Sinai and an electronic health record data pull at Geisinger. Patients were recruited by phone and/or email and patient interviews were conducted from 8/2023-12/2024. Patients recruited from Mount Sinai were purposively sampled for racial and ethnic diversity. Individuals at Mount Sinai whose primary language is Spanish were also prioritized.

For clinician interviews, participants included primary care providers, cardiology specialists (i.e., cardiologists, lipidologists), and genetic counselors at both institutions. As this study focuses on implementing genomic information into care broadly, we purposively sampled to include more clinicians working in primary care and internal medicine. Clinicians were recruited by email and interviews were conducted from 6/2023-4/2024. All clinicians were interviewed individually with the exception of two primary care providers who participated in an interview together.

Interviews were conducted via video-conference or phone with a deliberative engagement style where participants were presented with vignettes of hypothetical scenarios representing the causes of severe hypercholesterolemia: FH, high-risk PRS, elevated Lp(a), or environmental/lifestyle/behavioral factors (i.e., no known genetic etiology). Slides with the vignettes were emailed to participants to review before the interview. During the interviews, slides were shared with participants via screensharing on the video-conference call or verbally described if the interview was conducted by phone. Clinician and English-speaking patient interviews were conducted by experienced qualitative researchers (L.K.J., G.C.S., Z.M.S.). A bilingual research coordinator (L.C.) conducted the Spanish-speaking interviews after extensive training by an expert in qualitative methods and interviewing (G.C.S). Participants received a $50 gift card upon completion of their interview.

Patient and were an average of 43 and 54 minutes long, respectively. Interview questions, incorporating elements from CMIR^16^ focused on patient and clinician perspectives about how they would respond to each identified cause of severe hypercholesterolemia, their needs and preferences for communicating about and acting on these results, and barriers and facilitators to the identification and management of the underlying causes of severe hypercholesterolemia.

Participants responded to demographic questions at the end of the interview. Interviews were audio-recorded with verbal consent from participants. Audio-recordings were transcribed and de-identified, checked for accuracy, and analyzed by the study team. Transcripts were reviewed and discussed by the interview team to assess when theoretical sufficiency was reached across patient and clinician participants at both institutions. Theoretical sufficiency was determined when the richness of data in each participant group covered a breadth of concepts and themes and no new data addressing the research questions were emerging. Recruitment continued until theoretical sufficiency was reached and interviews that were already scheduled after this point were completed to further ensure theoretical sufficiency^20,21^.

Interviewers continually debriefed after interviews to discuss emergent patterns, refine probes, and ensure theoretical sufficiency. A codebook thematic analysis in a post-positivist paradigm was conducted to better understand the current state through the lens of an established framework, while simultaneously acknowledging the identification of emergent themes through participant perspectives^22,23^. Interviews were thematically analyzed using the constant comparative method and sensitizing to constructs from CMIR (i.e., acceptability, appropriateness, adoption, feasibility, maintenance). A codebook was developed to define and operationalize each sensitizing construct. Two authors (G.C.S, L.K.J.) with expertise in implementation science and qualitative methodologies conducted open coding of a random sample of three patient transcripts and three clinician transcripts to confirm participants’ reports aligned with CMIR concepts and iterated on the codebook. One study team member then coded each transcript as a primary coder (Z.M.S.) and a senior team member served as a secondary coder (G.C.S or M.L.G.H.). The primary coder independently coded each transcript using the codebook, with review by the secondary coder. Discrepancies were resolved by consensus with the rest of the coding team (G.C.S., Z.M.S., A.R.K., D.C., M.K., K.M.M, M.L.G.H.) and the codebook was revised to improve its clarity and consistency.

The coded data was segmented based on participant type (i.e., patient or clinician) and implementation outcome (i.e., acceptability, appropriateness, adoption, and feasibility) using Atlas.ti software version 23 to facilitate axial coding of the resulting four patient and four clinician sections of segmented data (e.g., patient acceptability, clinician acceptability). Each section was assigned to a member of the axial coding team (Z.M.S., M.K., K.M.M., M.L.G.H) to identify themes describing anticipated implementation outcomes of identifying and managing the underlying causes of severe hypercholesterolemia. Axial coding team members were sensitized to differences in the emerging themes based on severe hypercholesterolemia etiology, institution, clinician type, or primary patient language. Results for each section were iteratively discussed and refined with the full coding team. Final themes for each implementation outcome were compared to each other and consolidated within the patient and clinician results where similarities were identified. The contributing CMIR implementation outcomes associated with each overarching theme are noted in the result tables.

## Results

A total of 28 patients and 25 clinicians were interviewed across Geisinger [G] (13 Patients, 12 Clinicians) and Mount Sinai [MS] (15 Patients, 13 Clinicians). Among Mount Sinai patient interviews, 7/15 of these were conducted with patients whose primary language was Spanish [SP]. Interviewed clinicians included 11 primary care providers [PCP], seven cardiology specialists [C] and seven genetic counselors [GC]. Patient and clinician demographics are further described in Tables 1 and 2. The themes identified in the patient and clinician groups are described separately below. As described below, differences were observed based on severe hypercholesterolemia etiology, patient and clinician institution, and clinician type. Differences were not noted between patients with different primary languages (English or Spanish).

**Table 1.**
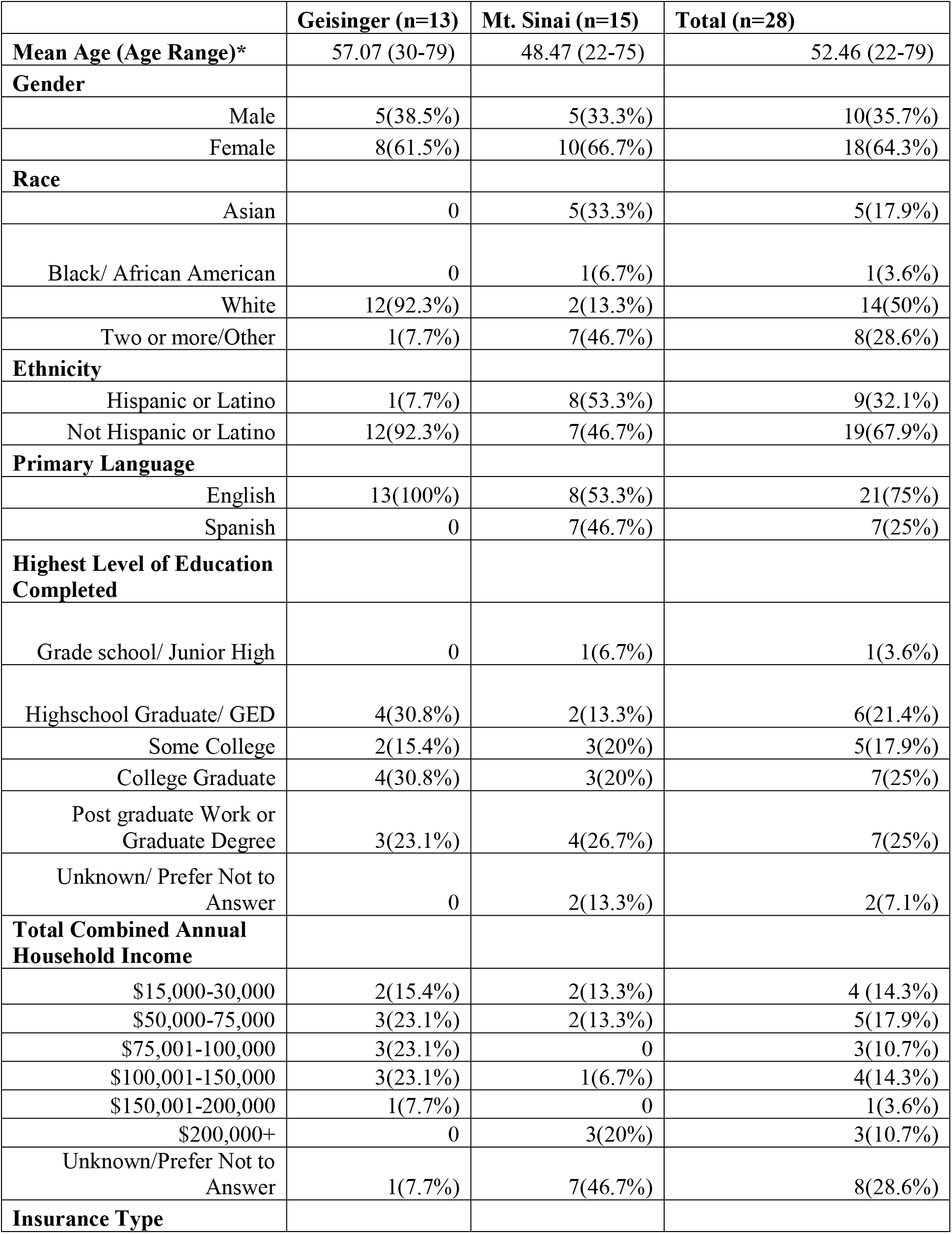
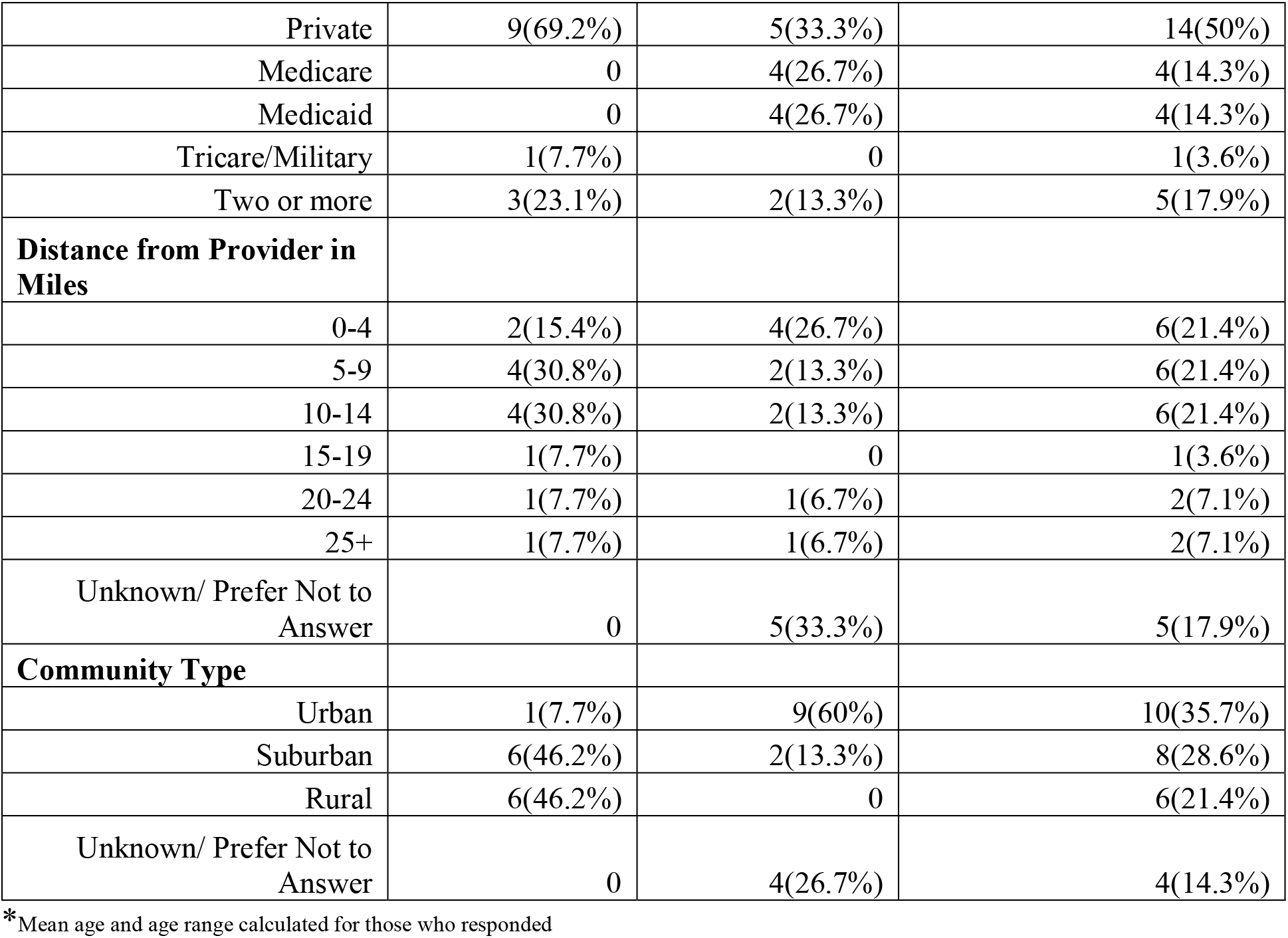
Patient Demographics.

|  | **Geisinger (n=13)** | **Mt. Sinai (n=15)** | **Total (n=28)** |
| --- | --- | --- | --- |
| **Mean Age (Age Range)\*** | 57.07 (30-79) | 48.47 (22-75) | 52.46 (22-79) |
| **Gender** |  |  |  |
| Male | 5(38.5%) | 5(33.3%) | 10(35.7%) |
| Female | 8(61.5%) | 10(66.7%) | 18(64.3%) |
| **Race** |  |  |  |
| Asian | 0 | 5(33.3%) | 5(17.9%) |
| Black/ African American | 0 | 1(6.7%) | 1(3.6%) |
| White | 12(92.3%) | 2(13.3%) | 14(50%) |
| Two or more/Other | 1(7.7%) | 7(46.7%) | 8(28.6%) |
| **Ethnicity** |  |  |  |
| Hispanic or Latino | 1(7.7%) | 8(53.3%) | 9(32.1%) |
| Not Hispanic or Latino | 12(92.3%) | 7(46.7%) | 19(67.9%) |
| **Primary Language** |  |  |  |
| English | 13(100%) | 8(53.3%) | 21(75%) |
| Spanish | 0 | 7(46.7%) | 7(25%) |
| **Highest Level of Education Completed** |  |  |  |
| Grade school/ Junior High | 0 | 1(6.7%) | 1(3.6%) |
| Highschool Graduate/ GED | 4(30.8%) | 2(13.3%) | 6(21.4%) |
| Some College | 2(15.4%) | 3(20%) | 5(17.9%) |
| College Graduate | 4(30.8%) | 3(20%) | 7(25%) |
| Post graduate Work or Graduate Degree | 3(23.1%) | 4(26.7%) | 7(25%) |
| Unknown/ Prefer Not to Answer | 0 | 2(13.3%) | 2(7.1%) |
| **Total Combined Annual Household Income** |  |  |  |
| \$15,000-30,000 | 2(15.4%) | 2(13.3%) | 4 (14.3%) |
| \$50,000-75,000 | 3(23.1%) | 2(13.3%) | 5(17.9%) |
| \$75,001-100,000 | 3(23.1%) | 0 | 3(10.7%) |
| \$100,001-150,000 | 3(23.1%) | 1(6.7%) | 4(14.3%) |
| \$150,001-200,000 | 1(7.7%) | 0 | 1(3.6%) |
| \$200,000+ | 0 | 3(20%) | 3(10.7%) |
| Unknown/Prefer Not to Answer | 1(7.7%) | 7(46.7%) | 8(28.6%) |
| **Insurance Type** |  |  |  |
| Private | 9(69.2%) | 5(33.3%) | 14(50%) |
| Medicare | 0 | 4(26.7%) | 4(14.3%) |
| Medicaid | 0 | 4(26.7%) | 4(14.3%) |
| Tricare/Military | 1(7.7%) | 0 | 1(3.6%) |
| Two or more | 3(23.1%) | 2(13.3%) | 5(17.9%) |
| **Distance from Provider in Miles** |  |  |  |
| 0-4 | 2(15.4%) | 4(26.7%) | 6(21.4%) |
| 5-9 | 4(30.8%) | 2(13.3%) | 6(21.4%) |
| 10-14 | 4(30.8%) | 2(13.3%) | 6(21.4%) |
| 15-19 | 1(7.7%) | 0 | 1(3.6%) |
| 20-24 | 1(7.7%) | 1(6.7%) | 2(7.1%) |
| 25+ | 1(7.7%) | 1(6.7%) | 2(7.1%) |
| Unknown/ Prefer Not to Answer | 0 | 5(33.3%) | 5(17.9%) |
| **Community Type** |  |  |  |
| Urban | 1(7.7%) | 9(60%) | 10(35.7%) |
| Suburban | 6(46.2%) | 2(13.3%) | 8(28.6%) |
| Rural | 6(46.2%) | 0 | 6(21.4%) |
| Unknown/ Prefer Not to Answer | 0 | 4(26.7%) | 4(14.3%) |

\*Mean age and age range calculated for those who responded

**Table 2.** Clinician Demographics.

|  | Geisinger (n=12) | Mt. Sinai (n=13) | Total (n=25) |
| --- | --- | --- | --- |
| **Clinician Type** |  |  |  |
| Primary Care Providers | 6(50%) | 5(38.4%) | 11(44%) |
| Genetic Counselors | 3(25%) | 4(30.8%) | 7(28.0%) |
| Cardiology Specialists | 3(25%) | 4(30.8%) | 7(28%) |
| **Mean Age (Age Range)\*** | 45.4 (26-68) | 43.5 (27-56) |  |
| **Gender** |  |  |  |
| Male | 6(50%) | 2(15.4%) | 8(32%) |
| Female | 5(41.7%) | 7(53.8%) | 12(48%) |
| Unknown/ Prefer Not to Answer | 1 (8.3%) | 4(30.8%) | 5(20%) |
| **Race** |  |  |  |
| Asian | 1 (8.3%) | 3(23.1%) | 4(16%) |
| White | 10(83.3%) | 5(38.5%) | 15(60%) |
| Two or More | 0 | 1(7.7%) | 1(4%) |
| Unknown/ Prefer Not to Answer | 1(8.3%) | 4(30.8%) | 5(20%) |
| **Ethnicity** |  |  |  |
| Not Hispanic or Latino | 11(91.7%) | 9 (69.2%) | 20 (80%) |
| Unknown/ Prefer Not to Answer | 1(8.3%) | 4(30.8%) | 5(20%) |
| **Years of Experience** |  |  |  |
| Under 5 | 3(25%) | 4(30.8%) | 7(28%) |
| 5-9 | 2(16.7%) | 0% | 2(8%) |
| 10-14 | 2(16.7%) | 1(7.7%) | 3(12%) |
| 15-19 | 0 | 2(15.4%) | 2(8%) |
| 20 or More | 4(33.3%) | 2(15.4%) | 6(24%) |
| Unknown/ Prefer Not to Answer | 1(8.3%) | 4(30.8%) | 5(20%) |
| **Years at Institution** |  |  |  |
| Under 5 | 4(33.3%) | 4(30.8%) | 8(32%) |
| 5-9 | 2(16.7%) | 1(7.7%) | 3(12%) |
| 10-14 | 4(33.3%) | 2(15.4%) | 6(24%) |
| 15-19 | 0 | 1(7.7%) | 1(4%) |
| 20 or More | 1(8.3%) | 1(7.7%) | 2(8%) |
| Unknown/ Prefer Not to Answer | 1(8.3%) | 4(30.8%) | 5(20%) |

\*Mean age and age range calculated for those who responded

### Patient Themes

Qualitative analysis of patient interviews revealed four themes describing patients’ preferences for learning about and managing results across severe hypercholesterolemia etiologies: 1) The importance of a conversation with their clinician about results and recommended care steps, 2) The helpfulness of credible take home resources, 3) Anticipated actionable care changes for all results with varying associated reactions, and 4) A desire for multidisciplinary care coordination and barriers to receiving this care (Table 3).

**Table 3.** Patient Themes.

| **Theme 1. Importance of a results conversation including recommended care steps (acceptability, feasibility, adoption)** |  |
| --- | --- |
| Varying communication preferences | *"I would like the doctor to explain it to me... because sometimes you can see information on paper, but you may not understand it the way doctors understand it" (5SP-MS)* |
|  | *"I would rather go in person to my PCP to talk about it more... I'll be more comfortable by in person talking to her than talking to her over the phone so I can comprehend more of what she's saying." (10P-G)* |
|  | *"For me, it's the combination of explaining to me over the phone, having the ability to ask questions and then send me the data so I can share with other doctors maybe or other family members or can research it by myself." (8P-MS)* |
| In-person support for Lp(a) and PRS | *"The doctor you're most familiar with, I think would be the best way to go and have a face-to-face talk and have this all [PRS] explained in layman's terms so that, you know, you could understand what they're talking about." (4P-G,)* |
|  | *"I don't even know what a lipoprotein is...I want to meet the doctor, see the doctor face to face and have him explain this to me in person. Even, even with props, if necessary." (6P-G)* |
|  | *"I think it's [PRS] less familiar to me than the other one. Give me the data, share with me like how I can do more research...I would like to understand better, like have maybe more examples." (8P-MS)* |
|  | *"I just feel like this one [Lp(a)] seems a little more serious for me...I'd rather I'd rather go in person and get everything done out there and get all the information out now" (4P-MS)* |
| Information about health risks, management, and family impact | *"What are your...risks for having a heart attack or stroke? ...I think probably would be the most important." (4P-G)* |
|  | *"The treatment options, pros and cons of each, how they would affect or not affect my lifestyle, and sort of a risk/benefit analysis of treating versus doing nothing...what is my...risk of a heart attack or stroke if I do nothing versus if I do something." (5P-MS)* |
|  | *"What are the chances that I have passed it on to anybody else? Who in family would be also likely to be affected...if there's any recommendations regarding notification...I would do that as well." (3P-MS)* |
| **Theme 2: Helpfulness of credible take home resources (feasibility, appropriateness, acceptability, adoption)** |  |
| Resources in various formats mirroring information from clinician | *"Links to website...a booklet from the clinic .. you could read more about it...I can actually go in and read about it... how to prevent that, what type of food to eat and how to lower your risk of getting heart attack." (2P-MS)* |
|  | *"I do like the idea of something I can read about. ... 'I've read the...pamphlets that you gave me, I watched the...video you gave me...I still have questions. Can you and I sit down and talk about this?' ...That would be helpful." (2P-G)* |
|  | *"Well, I do some more research on my own and then, you know, start following the guidelines as to what the, you know, what the doctor's laid out." (12P-G)* |
| Interest in doing further research on own time | *"I'm the kind of person that does my own research, I don't necessarily think getting a direct link to something is a bad idea. I think that's also helpful. You know, and that can help tailor my research."* (1P-MS) |
| **Theme 3: Identifying the cause of hypercholesterolemia can prompt actionable care changes with varying reactions to receiving this risk information (adoption, appropriateness, feasibility, acceptability)** |  |
| FH is important to know about for patients and their family | *"...it [FH] will be important mainly because it explains why –it makes the plan moving forward better...if it's like flagging other family members saying...you need to get checked...I think there's a difference in how to act on it...If it's genetic...then maybe intervention is more important."* (8P-MS) |
|  | *"Knowing what it [FH] is and that it is familial, I have a daughter... I will do anything to make sure she doesn't get her chest cracked...finding this out means that we have an incredibly high chance of not letting this happen to her."* (2P-G) |
| Lp(a) and PRS are informative but may prompt fear | *"For somebody like me that has gone through an MI...there's always a question of why and why me?...it is always a relief to have some understanding of what happened and why."* (3P-MS) |
|  | *"I think it [PRS] would be like more serious... I feel like I would be like way more distraught...I think I would be a little more concerned about it because there were so many gene mutations."* (6P-MS) |
|  | *"Extremely worried, scared... I think it's a more serious problem; it's very serious for someone to have something like that [Lp(a)] in their body regarding cholesterol"* (3SP-MS) |
| Continued follow-up if no genetic cause identified | *"It's not a, 'hey, I can put you on this medication and then it goes away.' This is an ongoing, 'we really have to keep an eye on it,' even if it's just once every 6 months we take these tests or whatever it happens to be. That's an ongoing thing."* (2P-G) |
| Not identifying a genetic cause may prompt relief or frustration | *"I think some patients or me can feel relieved at some level because they say it's not genetic. I have control of it. It's probably diet and exercise, right? Or...maybe also like a little bit frustrated...I don't know what's going on right now."* (8P-MS) |
| Impact of ancestry on access to actionable risk information | *"I did undergo some genetic testing, but the genetic testing was not conclusive because the genetic test I guess was more geared towards the European population"* (3P-MS) |
|  | *"I am of...South Asian descent. I got tested for FH and it was negative...my LDLs were way over 190s, and they didn't really have a great answer... it'd be nice to have more thorough answers for a specific ethnicity."* (1P-MS) |
| **Theme 4: Desire for multidisciplinary care and potential system-level barriers to receiving such care (adoption, feasibility, appropriateness)** |  |
| Access to specialists | *"I would like the doctor to refer me to a nutritionist, since that person should know more about diets than my doctor. That nutritionist will...give me all the necessary recommendations."* (4SP-MS) |
|  | *"...just like talking to like a therapist or social worker to make sure like, hey, it's OK, like although...we don't know a lot of information about this [Lp(a)], you're still gonna be OK."* (6P-MS) |
| The importance of expertise for PRS and Lp(a) | *"You tend to listen to your specialists a little bit more sometimes because you're not just a general, 'I'm taking care of all of you,' kind of a person. They are a very specific, 'this is your heart.' And you can lose your toes, but you can't lose your heart." (2P-G)* |
|  | *"I'd honestly wanna make sure that whoever's treating me as a clinician is kind of at the top of their... knowledge base for the disease process as well as someone that's kind of familiar with treating it" (1P-MS)* |
| A desire for PCP involvement at Geisinger | *"You know my family doctor...If there's anything going on with my cholesterol, she would probably notice it right away." (3P-G)* |
|  | *"I definitely would want my GP to be the one to talk to me about it. 'Cause that's the doctor you're going to be more comfortable with, you know, and is, you know, going to probably be a little more compassionate and understanding as she's he or she or he is explaining it to you." (4P-G)* |
| Potential barriers to follow-up care | *"That's a problem with the hospital...Sometimes you have a 15-minute visit, and it's terrible. they don't give you enough time, or they don't explain things in a way you can understand." (7SP-MS)* |
|  | *"They typically have a only a certain amount of time to have with you." (13P-G)* |
|  | *"Don't change my doctor. Every six months, they switch...I have to explain everything all over again...they ask me the same questions." (2SP-MS)* |
|  | *"Yeah, I think if I, if there's a barrier to entry, you're more likely to become complacent and not want to get things done. And I think of things like, OK, here's your next three-month lab draw order...If you make things a lot easier to get done in the first place...I think you're more likely to get those things done versus if you have to call back to the office, talk to your doctor, get the orders placed... you're not gonna get it done" (1P-MS)* |

Abbreviations: Mount Sinai (MS), Geisinger (G), Patient (P), Spanish Language Patient (SP), Primary Care provider (PCP)

First, patients emphasized the importance of a conversation with their clinician about their result and recommended next steps (Theme 1). While patients agreed that results disclosure by their clinician was important to ensure understanding (5SP-MS), their preferences about mode of communication varied. Some participants described in-person conversations as more comfortable and informative (10P-G), while others felt that an explanation over the phone was appropriate (8P-MS). For Lp(a) and PRS results specifically, patients were more interested in an in-person results discussion that included a clear explanation in care terms they understood (4P-G). Reasons for these preferences included less familiarity (6P-G; 8P-MS) and the perceived severity of these results (4P-MS). Across result types, patients sought information about their health risks, management guidelines, and how the result would impact their family during results disclosure. Patients described wanting to know how likely they would be to have a heart attack or stroke (4P-G), to understand the pros and cons of different treatment options (5P-MS), and to receive guidance about notifying at-risk family members (3P-MS).

Second, patients described the helpfulness of credible take home resources to accompany the results discussion with their clinician (Theme 2). Patients described how these resources in various formats (e.g., video, pamphlet, website) that mirrored the information shared during results disclosure could prompt them to learn more on their own time (2P-MS) and ask their clinician follow-up questions (2P-G). Additionally, participants described how they may wish to complete their own research before acting on their care plan (12P-G) and requested links from their clinician to facilitate this (1P-MS).

Next, patients anticipated that all result types could prompt actionable changes to their care, with varying reactions to receiving this risk information (Theme 3). Overall, patients welcomed information about FH, anticipating that these results could prompt intervention to improve health for themselves and their family members (2P-G, 8P-MS). For Lp(a) and PRS, some patients anticipated these results would be similarly informative and may help explain their health history (3P-MS). Participants also anticipated fear or concern related to Lp(a) and PRS, connecting this reaction to their perception of the severity of these results (SP3-MS; 6P-MS). If no genetic cause was identified (i.e., hypercholesterolemia due to environmental/lifestyle/behavioral factors), patients anticipated ongoing follow-up to continue to investigate underlying causes and manage their cholesterol (2P-G) and articulated either relief that nothing serious was identified or frustration that the cause of their high cholesterol couldn’t be explained (8P-MS). Importantly, several patients described their own experiences receiving inconclusive results related to limitations of genetic information outside of European ancestries and a desire for more tailored information to guide their care (3P-MS, 1P-MS). Of note, anticipated reactions to and feelings about the underlying causes of severe hypercholesterolemia were predominantly shared by patients at Mount Sinai, while patients at Geisinger did not discuss their anticipated emotional responses to these results in detail.

Last, patients described a desire for multidisciplinary care and noted potential system-level barriers to their follow-up care (Theme 4). Patients discussed the importance of access to specialists (e.g., cardiology, nutrition, therapists, social workers). They expressed that nutritionists could help patients make appropriate diet changes (4SP-MS) and how therapists or social workers could provide psychosocial support (6P-MS). Particularly for high Lp(a) and PRS result types, patients emphasized the importance of involving perceived experts in their care.

Patients explained how recommendations from cardiology may carry more weight than general clinicians (2P-G) and how they would like to be seen by clinicians who are knowledgeable and experienced with these result types (1P-MS). At Geisinger, some patients expressed an expectation for primary care involvement in their care due to their familiarity and established relationship with these clinicians (3P-G, 4P-G). Across systems, patients described how lack of care continuity and limited clinician time could negatively influence their access to care. Patients described that appointments were often too short to comprehensively discuss their results and recommended management (7SP-MS, 13P-G) or that their care team frequently changed, which could lead to gaps in their recommended management (2SP-MS). To streamline follow-up care, patients proposed scheduling out anticipated follow-up appointments and placing required orders further in advance (1P-MS).

### Clinician Themes

Qualitative analysis of the clinician interviews revealed four themes describing how clinicians would approach the identification and management of severe hypercholesterolemia etiologies: 1) The importance of integration with existing electronic health record (EHR) workflows, 2) Limited understanding of certain causes of severe hypercholesterolemia, particularly Lp(a) and PRS, based on stage of clinical implementation and their own clinical experiences, 3) LDL-C levels and multidisciplinary care are key to management, and 4) Tailoring care to the individual patient (Table 4).

**Table 4.** Clinician Themes.

| **Theme 1: Importance of integrating test ordering and follow-up into existing EHR workflows (feasibility, acceptability, appropriateness)** |  |
| --- | --- |
| Current EHR navigation challenges | *"If it comes from somewhere else...it's harder for me to get that to the patient. Some genetic tests come through the results section, but some of them don't, and so it's not consistent. And that I think that makes it challenging... disturbs... regular workflow and the way I do things." (5PCP-MS)* |
|  | *"This is in general, not just for cholesterol, but I have to often go searching back. What did the cardiologist say about this? Or what did the pulmonologist say about this? Whereas, if it was documented clearly in that condition...then everybody could see it and access it." (7PCP-G)* |
|  | *"...ideally like a way to integrate reports into electronic medical records. I think is ideal and increasingly...labs are starting to be able to integrate them so that they show up in the labs section of Epic or other EMRs." (1GC-MS)* |
| Request for succinct results communication through the EHR | *"I think that we're in the realm of, of the in-basket we call it. It's just where results come in and it works...It really works because the next thing you can do is right away contact the patient." (8C-G)* |
|  | *"I think most patients want to know what their risk of either cardiovascular event or even death... whatever predictive information that...I can convey safely to a patient...the stepwise action plan, that would be how I want that." (6C-MS)* |
|  | *"So, I think having just like the biggest few key points, which ultimately for a lot of people will lead to more questions, but that's the purpose of the meeting with providers... I think rather than having the patient Google just providing them with a little handout with the big points would be helpful." (10GC-G)* |
| Request for clear communication in the EHR between clinicians | *"I do appreciate the direct communication with a note from the provider or like a cc, a routing through the EMR, hey, we've done this work, and this is what we found." (7PCP-MS)* |
|  | *"I'm not necessarily going to like it, but you need to be very clear that you're telling me that it's my job and my responsibility to deal with it...people play hot potato with stuff if they're not sure about it...if I get the message, it better be like, 'Hey this is on you to talk to the patient about' ...primary care is there to discuss cholesterol issues." (4PCP-G)* |
| **Theme 2: Different levels of understanding between severe hypercholesterolemia etiologies depending on the stage of clinical implementation and clinical experiences (acceptability, appropriateness, feasibility, adoption)** |  |
| Heterogeneity in current FH workflows | *"So, if I had ordered a test and I got a result that said they had FH, I guess if it was something that I had ordered, then I am probably punting that and sending that to our lipid clinic or to our cardiology team." (2PCP-G)* |
|  | *"FH is a condition, which I think in some cases it's appropriate for like a primary care provider to order testing, and it may not be [appropriate at times] ... You want that information...readily visible for a non-genetics provider who's ordering the testing for them to actually understand the significance of the result." (1GC-MS).* |
|  | *"My approach is to see if I could get them to do the genetic testing because they're sitting there in front of me right now. Send it off, simultaneously refer to genetics, and then go from there. -Some [attendings] would rather just...have them meet with the genetic counselors. I think there's like different levels of comfort." (6C-MS).* |
| Comfort with Lp(a) or uncertainty about the clinical utility | *"So if we're concerned, we can add this on to a lipid panel... this is something that we would, I could check in clinic, and we have done for patients...I think this triggers, I think this would trigger me to be quite aggressive in terms of wanting to start a patient on medication...and encouragement for the patient to do genetic testing." (6C-MS)* |
|  | *"I don't know what the additional value of this is. I mean on top of having a lipid profile like what added value does a lipoprotein(a) lab value have? Like what is it?" (10PCP-MS)* |
|  | *"It's not something that I was exposed to like in my training...what is Lp(a)? I think providing more information to the genetic counselor, seeing the patient about what this is, what this means, what we know, what we don't know is important." (11GC-G)* |
| PRS is not standard of care yet | *"I think the differences are around actionability first of all, because a monogenic risk is something we, have guidelines and parameters around and polygenic risk is still newer and not... medical recommendations aren't really guideline driven at this point, so that would be a big area of counseling." (13GC-MS)* |
|  | *"When you start getting into polygenic and something where it's less clear that it's an inherited condition, is there's a lot of hedging and hawing and a lot of complicated stuff that the provider may not understand, that is then expected to be conveyed to the patient. This will become more part of routine care...but I don't think we're there yet." (4PCP-MS)* |
|  | *"The polygenic is kind of more of a gray zone...I would definitely want more...statistics to justify treatment 'cause that's the bottom line: Am I gonna treat them or not?...What are the guidelines today for treating patients?" (3PCP-G)* |
| Hypercholesterolemia due to environment/lifestyle/behavioral factors is routine in primary care | *"The genetic counselor sending me an FYI: "Hey, we got these results. Your patient has high cholesterol, but it's not a genetic issue." That's my bread and butter. This is what I do on a day-to-day basis... I just run with that, look at their other risk factors...reach out to the patient." (2PCP-G)* |
|  | *"This is what I'm doing every day, right? I'm sending a lipid panel on most people who I care for either because they're...of age for screening or because...they're on statin, and I'm checking them annually." (7PCP-MS)* |
| **Theme 3: The importance of LDL-C levels and the value of specialist involvement across severe hypercholesterolemia etiologies (feasibility, adoption, appropriateness)** |  |
| LDL-C guides patient management across etiologies | *"[I'm] not going to play around with Lp(a) - I'm going to go right to the LDL cholesterol and see what's happening there and do what is necessary to get those numbers to goal." (5PCP-G).* |
|  | *"If it's the LDL is usually greater than 190, then for me that's like a slam dunk of starting statins." (5C-MS)* |
|  | *"I think the starting point is same protocol from our standpoint, right...a little blurb saying, 'Look. This is associated...with, you know, higher risks, higher LDL-C.' And perhaps on the back end...have a reflex for LDL-C, that might useful." (1PCP-G)* |
| Multidisciplinary care coordination promotes patient management | *"I usually do have some cardiologists...I would refer them to. To, you know, just have a longer conversation...They're coming back to talk to me about it, but if they haven't gotten a chance to speak to a genetic counselor...that's another resource that we have that I could have someone talk to." (2PCP-MS)* |
|  | *"...as a genetic counselor not being able to prescribe medications myself or order imaging tests...making sure that they have a specialist who is familiar with hypercholesterolemia and how Lp(a) plays a role, getting handed off to that cardiologist or lipidologist would be super important." (13GC-MS)* |
|  | *"There's all kinds of things that we that we can do as a team approach, including cardiovascular, pharmacists...I look at the whole cholesterol thing as being a team approach...I think we need to...continue to do it that way." (12C-G)* |
| **Theme 4: Tailoring care to the individual patient (adoption)** |  |
| Cultural adaptations to their results communication and resources at Mount Sinai | *"...in some cases it may not be appropriate to apply a polygenic risk score that was validated in like a white European population to a patient of Asian ancestry...I think it's really important that it be clear on the report which ancestry is the population was validated for." (1GC-MS)* |
|  | *"It needs to be multilingual, so that's really, really important... in at least English and Spanish, and that when you're talking about diet recommendations...the nutritionists are well aware of cultural differences in diet...and can adapt the recommendations to what is gonna feel right for someone culturally." (2PCP-MS)* |
| Different preferences for which clinicians are involved with care at Geisinger | *"...while I trust that the genetic counselors are gonna reach out and do a much better job with education for their patient than I ever would, sometimes the patients do count on us as their primary to answer their questions." (2PCP-G).* |
|  | *"I think I had one [patient] that basically said, 'I don't want anything, I just want to talk to my doctor about it,' and then that came back to me and I had walked her through it the next time she came in for a visit and tried to convince her to go see cardiology because I felt she needed it." (7PCP-G).* |

Abbreviations: Electronic Medical Record (EHR), Mount Sinai (MS), Geisinger (G), Primary Care Provider (PCP), Cardiology Specialist (C), Genetic Counselor (GC)

First, clinicians described the importance of embedding test ordering and follow-up into existing EHR workflows (Theme 1). They reflected on how current EHR workflows posed challenges. Clinicians explained that genetic test results may be documented in different places depending on how they were ordered (5PCP-MS) and the information required to manage patients was often spread across the EHR (7PCP-G). They recommended integration between external genetic test labs and the EHR and condition-specific documentation that included all information required for management in one place (1GC-MS, 7PCP-G). Clinicians requested EHR notifications (e.g., in-basket messages) about test results (8C-G) including a stepwise action plan (6C-MS) and key points for clinicians to address with patients (10GC-G). Further, clinicians highlighted the importance of communication between clinicians within the EHR.

Primary care providers explained that when testing was ordered by another clinician, it was important for them to receive a summary of what was found (7PCP-MS) and, if they were expected to manage the result, directive information about this responsibility (4PCP-G).

Next, clinicians expressed different levels of uncertainty between causes of severe hypercholesterolemia depending on the stage of clinical implementation and their own clinical experiences (Theme 2). For FH, clinicians identified heterogeneity in current workflows. While FH was usually addressed in specialty care (2PCP-G), some clinicians discussed it may be appropriate for non-genetics clinicians to order FH genetic tests on a case-by-case basis (1GC-MS). Within specialty care, cardiologists explained that some were comfortable with ordering testing and referring to genetics for results follow-up while others preferred to refer to genetics to order testing (6C-MS). For Lp(a), the subset of clinicians who routinely ordered this testing (primarily cardiology specialists) described how these results could influence how urgently or aggressively they counseled patients about medical management (6C-MS), while clinicians who were unfamiliar with this result type were unsure of its clinical utility (10PCP-MS) and desired education about it (11GC-G). Clinicians agreed that PRS was not currently standard of care.

They viewed this result type as more complex for patients and clinicians with a need for additional evidence to guide clinical care (13GC-MS, 4PCP-MS, 3PCP-G). Severe hypercholesterolemia without an identified genetic cause was viewed as the most commonly encountered clinical scenario by all clinician types. Primary care providers described how they regularly counseled patients with hypercholesterolemia due to environment/lifestyle/behavior and were comfortable doing so (2PCP-G, 7PCP-MS).

Across result types, clinicians described how management decisions were driven by patient’s LDL-C and the importance of multidisciplinary care to achieve management goals (Theme 3). Clinicians explained that receiving results about the cause of patients’ severe hypercholesterolemia would prompt them to monitor and manage patient’s LDL-C (5PCP-G) and how they would recommend lipid lowering therapy based on patients’ LDL-C levels (5C-MS). Further, they suggested that management decisions could be facilitated if an LDL-C level were reflexively ordered once a relevant form of severe hypercholesterolemia were identified (1PCP-G). Clinicians explained how multidisciplinary teams promoted effective management. Primary care providers described that referrals to genetic counseling provided opportunities for longer conversations with experts and a second opinion when needed (2PCP-MS). Genetic counselors explained the limits of their own scope of practice and the importance of a handoff to a clinician well versed in hypercholesterolemia to prescribe medications and order any recommended follow-up tests (13GC-MS). Cardiology specialists echoed the importance of a team approach to cholesterol management, for example when describing their involvement of pharmacists in such care (12C-G).

Last, clinicians described how the care they provided was tailored to the individual patient (Theme 4). Clinicians highlighted the importance of patients’ ancestry and culture. When discussing PRS specifically, they described how the performance of the test may vary based on patients’ ancestry and the importance of clear information for which populations the testing has been validated (1GC-MS). When describing patient education materials, clinicians recommended having materials available in multiple languages and cultural adaptations within the materials, for example accounting for cultural differences in diet when providing nutrition recommendations (2PCP-MS). Additionally, clinicians acknowledged that patients may have different preferences for which clinicians are involved with their follow-up care. Geisinger primary care providers described how some patients turn to them as their primary source for recommendations and explained their role in aiming to meet patients’ informational needs (2PCP-G) and discussing with patients when and why they should consider seeing a specialist (7PCP-G).

## Discussion

While personalized risk information provides opportunities for cardiovascular disease prevention, evidence about the efficacy of current cardiovascular risk scores is modest^24^.

Incorporating genomics (e.g., FH, PRS, and Lp(a)) can increase the accuracy and resulting value of these risk scores in unaffected populations. This study evaluated patient and clinician perspectives about identifying and managing FH, high-risk PRS, elevated Lp(a), and severe hypercholesterolemia without a known genetic cause in clinical care. Importantly, this evaluation incorporated perspectives from multiple clinician types involved in hypercholesterolemia care at two distinct institutions. Patients and clinicians across institutions shared complementary perspectives that can inform the potential future implementation of comprehensive genomically informed ASCVD risk stratification.

Patients and clinicians saw value in understanding the causes of severe hypercholesterolemia and how these results inform cholesterol management. They desired clear information about the associated medical risks, recommended management, and familial impact for all result types. To promote actionability, patients requested take-home resources mirroring the information from their clinician and clinicians emphasized the importance of patients LDL-C levels to guide management. Given the prevalence of high cholesterol, there are numerous cholesterol patient education materials that are publicly available online. A recent analysis demonstrated an average Flesch-Kincaid Grade Reading level of 11^th^ grade among these materials^25^. Taken together with patients’ desire to do their own research, there is a need to ensure accessible resources for severe hypercholesterolemia are available. While clinicians acknowledged the differences in severe hypercholesterolemia result types, their request for LDL-C levels to guide management across results aligns with how currently available dyslipidemia guidelines are structured (e.g., LDL-C treatment goals based on ASCVD risk) ^3^. LDL-C based management recommendations and available information on how the cause of severe hypercholesterolemia may influence this should be incorporated into patient and clinician-facing materials to promote shared decision making.

Patients and clinicians agreed about the involvement of specialists in hypercholesterolemia management as a key facilitator. Patients described how the expertise provided by cardiology, nutritionists, therapists, and social workers could benefit their care, while clinicians explained the importance of referrals to genetic counseling, cardiology, and pharmacy to ensure patients’ needs are met. One established approach to specialist involvement in hypercholesterolemia care is multidisciplinary lipid management^26,27^. While such care can improve cardiovascular health, the required health system resources may limit access. Strategies to improve access, including pharmacist-led models and telemedicine visit types, are being explored^27,28^. We expected to see more differences between the urban and rural settings of our study population due to previously cited disparities in access to care and cardiovascular outcomes between urban and rural communities, however participants across systems shared similar experiences^29,30^. An exception was an emphasis on the continued involvement of primary care expressed by patients at Geisinger. While this is likely related to the central role of primary care in Geisinger’s integrated care model, it is also consistent with national trends among the general public demonstrating a preference for primary care to manage their health concerns^31,32^. Given the central role of primary care providers in lipid screening in the general population, effective approaches to longitudinal care coordination between specialists and primary care are needed.

Patients and clinicians explained how their perspectives were influenced by the underlying causes of severe hypercholesterolemia. A higher level of support was requested for Lp(a) and PRS as participants described uncertainty due to less knowledge, lower familiarity, and fewer medical guidelines for care decisions in these scenarios. Lp(a) remains underutilized in clinical practice despite recent guidelines^10,11^, however, we found that the subset of clinicians who ordered this testing regularly as early-adopters (e.g., cardiologists and lipidologists), were more comfortable doing so, suggesting that continued exposure to this testing in clinical practice may help address current perceptions about its utility^33,34^. Clinician perspectives that PRS is not yet standard of care aligns with current guidelines and challenges, including clinician knowledge, insurance coverage, and applicability in diverse populations^3,35^. Patients also raised concerns about the impact of their ancestry on access to accurate genomic risk information, further highlighting this limitation of PRS, which are derived predominantly from European-ancestry cohorts and lose predictive performance when applied to other populations^36^. High interest in PRS among clinicians in this study and others^37^ warrants future work to address current barriers. Our findings suggest that patient- and clinician-facing PRS resources should make ancestral applicability explicit rather than leaving it as an unstated caveat.

Patients’ anticipated emotional reactions to high Lp(a) and PRS were predominantly shared by patients at Mount Sinai. While this may stem from potential cultural or social differences between sites, it also aligns with published research. Studies evaluating patients’ experiences receiving elevated Lp(a) results found that 30-40% of patients reported concern or anxiety upon result receipt^38,39^. Further, in a study evaluating patients’ reaction to receiving PRS results for multiple complex disorders (e.g., type 2 diabetes, breast cancer) in a direct-to-consumer setting, 60% of participants reported a negative reaction in response to their results^40^. These studies suggested the need for enhanced patient support and resources, consistent with our findings about patients’ desire for in-person disclosure and emphasis on the importance of specialist involvement with these result types. Importantly, longer term follow-up with patients who have undergone such testing does not indicate sustained negative psychosocial impacts^41^. Lastly, while clinicians were comfortable with severe hypercholesterolemia due to environmental/lifestyle/behavior factors, patients reported guilt and uncertainty related to this result type, highlighting that patient support is beneficial even when a genetic cause is not identified^42^.

System-level barriers to caring for patients with chronic conditions including hypercholesterolemia are established as influential determinants of patients’ health outcomes^43^. In our study, patients highlighted limited time with clinicians and lack of continuity of care as challenging while clinicians described the need to establish clear care pathways that are integrated with current EHR workflows across result types. An analogous study evaluating patient and clinician perspectives about an automated intervention facilitating high-intensity statin prescriptions and lipid panels for patients with severe hypercholesterolemia, found that most patients filled the prescription recommended by their clinician and upstream barriers related to clinician awareness about the program and ineffective EHR workflows – both too many messages, and the lack of actionable alerts – posed implementation challenges^44^. While effectively integrated EHR workflows may help ameliorate some of the system-level barriers highlighted in our study related to care continuity and efficiency, these changes alone are insufficient and should be implemented with additional support, such as clinician education and audit and feedback, which have been demonstrated to improve the implementation of evidence-based practices^45^. Overall, multi-level implementation strategies that consider health system resource constraints (e.g., clinician time) highlighted in our study and others are needed to promote equitable access to care^46,47^.

There are a few limitations present in the current study. First, demographic information was not available for five (20%) of the clinicians interviewed. Among the available information, interviewed clinicians represented a wide range of years of clinical experience and specialties (e.g., primary care, genetic counselors), however the population predominantly self-reported as White, limiting the representation of perspectives from clinicians from different demographic backgrounds. While our patient sample was diverse in terms of race, ethnicity, educational attainment, income level, and insurance payors, including Medicare/Medicaid, this study only recruited from two healthcare institutions in the northeastern United States with active genomics research and clinical cardiovascular genetics services. Other healthcare systems, including those with less experience with genomics or those in different regions, may face different institutional barriers. Notably, Mount Sinai and Geisinger serve urban and rural settings, respectively, which enhances transferability of findings.

We sought to define the barriers and facilitators associated with the implementation of a complex intervention with multiple components (e.g., FH, PRS) that span different stages of clinical implementation. Perspectives from patients and clinicians suggest that to incorporate genetic information into ASCVD risk counselling the following criteria should be met: dedicated disclosure discussions accompanied by resources with actionable information for patients and clinicians, care coordination between primary and specialty care to support ongoing management, integration of resources and workflows within the EHR, and the development of clear implementation protocols to promote adoption. Further, although determinants recurred across etiologies, the support patients and clinicians sought varied based on the nature of the result, with a higher level of support (e.g., more education, additional specialist involvement) requested for probabilistic polygenic score (PRS) and quantitative heritable biomarker (Lp(a)) result types, which are at earlier stages of clinical implementation and were viewed with more uncertainty by patients and clinicians, compared to a deterministic monogenic variant (FH) or the absence of an identified genetic cause. Overall, participants desired genomic information to refine risk stratification for severe hypercholesterolemia, valued these results to guide care, and highlighted the need for improved knowledge across patients and clinician types to guide care management and support patients and families with severe hypercholesterolemia.

## Author Contributions

**Conceptualization:** LKJ, GCS, MLGH **Methodology:** LKJ, GCS, MLGH **Formal Analysis:** LKJ, GCS, ZMS, ARK, DC, MK, KMM, MLGH **Investigation:** KMM, GCS, ZMS, ARK, MIT, TN, MK, LC, DC, LKJ, MLGH **Writing – Original Draft:** KMM, GCS **Writing – Reviewing and Editing:** KMM, GCS, ZMS, MK, DC, LCa, LC, SSG, EEK, ARK, TN, MTO, VP, SS, MIT, LKJ, MLGH **Supervision:** GCS, LKJ, ARK, MLGH **Project Administration:** DC, LCa **Funding Acquisition:** GCS, SSG, MTO, LKJ, EEK

## Data Availability

Data supporting the findings reported in this manuscript is available from the corresponding author upon request to protect patient privacy.

## Acknowledgements

We thank all patients and clinicians who participated in this study for their time and the feedback they shared with us.

